# Prevalence and Factors Associated with Diabetes and hypertension Among HIV Patients at Tertiary Hospital Kilimanjaro Tanzania

**DOI:** 10.1101/2025.06.02.25328843

**Authors:** Agreyson L. Hindanya, George Kifumase, Joshua Baysa, Erick Mlaponi, Janeth J. Sabuni, Balthazar M. Nyombi, Isaack Lyaruu, Florida Muro, Dhahiri Mnzava, Debora C. Kajeguka

## Abstract

**Introduction:** Diabetes and hypertension are prevalent non-communicable diseases that significantly affect people living with HIV, particularly those on antiretroviral therapy. Despite the increasing recognition of these conditions, their prevalence and associated risk factors among people living with HIV in Tanzania remain underexplored. This study aimed to determine the prevalence and risk factors of diabetes and hypertension among HIV patients attending Child-Centred Family Care Clinic at KCMC Hospital.

**Methods:** A cross-sectional study was conducted from May to June 2024, involving 341 people living with HIV attending the Child-Centred Family Care Clinic at KCMC Hospital. Data were collected through face-to-face interviews, file reviews, and physical measurements. Hypertension was defined as systolic blood pressure ≥ 140 mmHg/ or diastolic blood pressure ≥ 90 mmHg. Diabetes was defined as blood glucose level of ≥140 mg/dL. A logistic regression model was used to identify factors associated with diabetes and hypertension.

**Results:** Prevalence of hypertension was 23.5% (80) while the prevalence of 14.1% (48) was diabetes. Hypertension was most prevalent in individuals aged 36-62 years (71.3%), whereas 70.9% of diabetic patients reported a family history of diabetes. Multivariate analysis revealed that older age was significantly associated with hypertension, with participants aged 18-35 years showing a lower likelihood of hypertension (AOR=0.10; 95% CI: 0.03-0.41; p=0.004) compared to those over 63 years. Participants earning TZs 500,000 or more per month were more likely to develop diabetes as compared to others (AOR=4.112; 95% CI: 1.199-14.108; p=0.025).

**Conclusion:** The findings indicate that socioeconomic determinants are critical in influencing the prevalence of diabetes and hypertension among people living with HIV, suggesting that interventions should focus on improving screening and early treatment to at risk groups.

## INTRODUCTION

HIV/AIDS remains a global epidemic, with approximately 39 million individuals infected worldwide as of 2022. The increasing prevalence of non-communicable diseases (NCDs) among people living with HIV (PLWH) has emerged as a significant public health concern, particularly in low- and middle-income countries(Patel *et al*., 2018).NCDs, including diabetes and hypertension, have been identified as major contributors to morbidity and mortality in PLWH (Badacho and Mahomed, 2023),(Hadavandsiri *et al*., 2023). In sub-Saharan Africa, the prevalence of NCDs among PLWH is estimated to be around 20.1%, highlighting the urgent need for comprehensive healthcare strategies that address both HIV management and the rising incidence of NCDs (Moyo-Chilufya *et al*., 2023). The prevalence of diabetes among PLWH varies widely across different studies; however, estimates suggest that it ranges from 6.8% to 26% in low- and middle-income countries (Sarkar and Brown, 2021). In Tanzania specifically, recent studies have reported a diabetes prevalence of approximately 13% among HIV-infected individuals (Jeremiah *et al*., 2020a).Hypertension prevalence among PLWH has been reported to be between 25% and 70% in various studies across sub-Saharan Africa (Isaac Derick and Khan, 2023).

The intersection of HIV and NCDs is complex, influenced by various factors such as socioeconomic status, access to healthcare, and the long-term effects of antiretroviral therapy (ART) (Moyo-Chilufya *et al*., 2023),(UNAIDS, 2019).Research indicates that PLWH are at an elevated risk for developing diabetes due to several factors, including lifestyle changes associated with HIV treatment and the metabolic effects of ART (Umar and Naidoo, 2021).

The management of diabetes and hypertension in PLWH requires a multifaceted approach that includes regular screening, patient education on lifestyle modifications, and appropriate pharmacological interventions. However, many healthcare systems in low-resource settings face significant challenges in providing comprehensive care for NCDs alongside HIV treatment.

Among other challenges that healthcare systems face includes limited access to diagnostic tools, and inadequate training for healthcare providers on managing co-morbidities (Getahun *et al*., 2020). There is a need to explore the factors in population living with HIV who are at increased risk for conditions such as diabetes and hypertension. The knowledge is crucial for developing effective interventions aimed at improving health outcomes for PLWH. Understanding these factors can inform targeted interventions that address the unique needs of this population (Badacho and Mahomed, 2023).

## METHODS

### Study design and setting

A cross-sectional study was conducted at Child-Centered Family Care Clinic (CCFCC) in KCMC hospital. KCMC hospital is a tertiary referral hospital in Tanzania located on the foothills of Mt Kilimanjaro and serving more than 15 million people in Northern Tanzania. KCMC has three dedicated HIV clinics, namely Care and Treatment Clinic (CTC), Child-Centered Family Care Clinic (CCFCC), and the Infectious Disease Clinic (IDC). Approximately 2630 HIV patients receive care and treatment in these clinics in a month. Patients attend clinics three days in a week, Monday and Thursday for adult at CTC and children at CCFCC then Friday only CTC for adult also there is special clinic during Saturday incudes adolescence and youth only.

### Study population

The study included PLHIV who attended clinics at the CCFCC. Participants with pre-existing conditions that could influence blood glucose or blood pressure levels—such as kidney disease, thyroid disease, cancer, and those who are pregnant or breastfeeding—were excluded from the study. Additionally, individuals unable or unwilling to provide the necessary information or blood samples were also excluded.

### Sample size calculation

The following formula was used to calculate the sample size,) Z^2^P(1-P)/E^2^. In this equation, n represents the required minimum sample size, Z denotes the degree of confidence, E signifies the desired precision, and PP indicates the estimated proportion of the population. For this study, a confidence level of 95% was selected, corresponding to a Z value of 1.96. The estimated proportion of individuals living with diabetes among people living with HIV was set at 0.28 (Hertz *et al*., 2022) based on prior studies. The desired precision was established at 0.05.

### Data collection

Data collection involved a multi-faceted approach utilizing face-to-face interviews, medical file reviews, and physical measurements. A pretested structured questionnaire was developed to gather information on socio-demographic characteristics, medical history, lifestyle factors, and family history of diabetes and hypertension. The questionnaire was validated through a pilot study involving 30 participants who were not included in the main study. The data collected from medical files included information regarding clinical conditions, co-morbidities, viral load results.

Physical measurements were taken by trained research assistants. Blood pressure was measured using a calibrated sphygmomanometer, with participants in a seated position after resting for at least five minutes. Hypertension was defined as having a systolic blood pressure of 140 mmHg or higher, or a diastolic blood pressure of 90 mmHg or higher. Diabetes was assessed through fasting blood glucose levels, with diabetes defined as a fasting glucose level of 126 mg/dL or higher or a previous diagnosis of diabetes.

### Statistical analysis

Data analysis was performed using SPSS version 25. Descriptive statistics were computed to summarize participant characteristics and prevalence rates of diabetes and hypertension. Bivariate analyses were conducted to explore associations between demographic variables and the outcomes of interest using chi-square tests. Logistic regression models were employed to identify factors significantly associated with diabetes and hypertension while controlling for potential confounders. Adjusted odds ratios (AOR) with 95% confidence intervals (CI) were calculated to quantify the strength of associations.

### Ethical consideration

Ethical approval to conduct this study was obtained from Kilimanjaro Christian Medical University College (KCMUCo) ethical review board with certificate number CRERC No.UG116/2024. Permission to conduct the study was obtained from the head of CCFCC at KCMC hospital. Written informed consent was obtained from all participants. Assent involved participants in the age range less or equals to17 years, who were legally not able to consent by themselves. Parents or guardians consented on behalf of participants who were under 18 years.

Participant’s information was kept confidential and only participants numbers was used in the questionnaire.

## RESULTS

### Socio-demographic characteristics

A total of 341 study participant were recruited. Majority 66.6% (227) were female, their mean age was 44.7, STD 17.1 ranging from 8-82, most of them their age ranges from 36-62 years 55.7% (190). Participant with primary education were mostly represented 44.6% (152). Most participants 52.2% (178) were self-employed and those with an income below TZs 50,000/= per month 42.8% (146) were more represented, whereby 85% (290) had number of people per household between 1-5 and most of them were dependents 67.2% (229), Table 1.

**Table 1:** Socio-demographic characteristics of the studied population (n=341)

| Variable |  | Frequency | Percent |
| --- | --- | --- | --- |
| Mean 44.7, STD = 17.1 |  |  |  |
| **Age** | 8-35 | 97 | 28.4 |
|  | 36-62 | 190 | 55.7 |
|  | >63 | 54 | 15.8 |
| **Gender** | Female | 227 | 66.6 |
|  | Male | 114 | 33.4 |
| **Level of education** | No formal education | 6 | 1.8 |
|  | Primary education | 152 | 44.6 |
|  | Secondary education | 143 | 41.9 |
|  | Tertiary education | 40 | 11.7 |
| **Employment status** | Unemployed | 31 | 9.1 |
|  | Employed | 54 | 17.0 |
|  | Self-employed | 178 | 52.2 |
|  | Student | 49 | 14.4 |
|  | Retired | 24 | 7.0 |
| **Income in TZs** | <50000 | 146 | 42.8 |
|  | 50000-200000 | 131 | 38.4 |
|  | 200000-500000 | 46 | 13.5 |
|  | >500000 | 18 | 5.3 |
| **Number of people per household** | 1-5 | 290 | 85.0 |
|  | 6-10 | 50 | 14.7 |
|  | >10 | 1 | 0.3 |
| **Dependable** | No | 112 | 32.8 |
|  | Yes | 229 | 67.2 |

1USD = 2685.68 TZs

### Prevalence of Diabetes among HIV patients

Out of 341 HIV patients, 14.1% (48) were diabetic, Figure 1. The prevalence of diabetes was notably higher among individuals aged 36 to 62 years, with a rate of 60.4% (29), compared to other age groups p<0.001. Among diabetic positive patients approximately 72.9% (35) were female. Most of the patients were dependants 70.8% (34). Fifty percent (24) had primary education level and in terms of employment status 43.8% (21) were self-employed, p=0.001. A significant majority of the subjects had a family history of diabetes, accounting for 70.9% (34) p<0.001. Most participants 66.7% (32) were using ART for the duration between 21-37 years p=0.003, Table 2.

**Table 2:** Association of demographic characteristics with Diabetes among HIV patients (n=341)

| Variable |  | Diabetic status |  | P value |
| --- | --- | --- | --- | --- |
|  |  | Non-diabetic<br>n (%) | Diabetic<br>n (%) |  |
| Age | 8-35 | 96 (32.8) | 1 (2.1) | *<0.001 |
|  | 36-62 | 161 (54.9) | 29 (60.4) |  |
|  | >63 | 36 (12.3) | 18 (37.5) |  |
| Gender | Female | 192 (65.5) | 35 (72.9) | 0.31 |
|  | Male | 101 (34.5) | 13 (27.1) |  |
| Education level | No formal education | 6 (2.1) | 0 (0.0) | 0.32 |
|  | Primary education | 128 (43.7) | 24 (50.0) |  |
|  | Secondary education | 127 (43.3) | 16 (33.3) |  |
|  | Tertiary education | 32 (10.9) | 8 (16.7) |  |
| Employment status | Un-employed | 21 (7.2) | 10 (20.8) | *0.001 |
|  | Employed | 49 (16.7) | 9 (18.8) |  |
|  | Self-employed | 157 (53.6) | 21 (43.8) |  |
|  | Student | 48 (16.4) | 1 (2.1) |  |
|  | Retired | 17 (5.8) | 7 (14.6) |  |
| Income in TZs | 200,000-500,000/= | 40 (13.7) | 6 (12.5) | 0.12 |
|  | 50,000-200,000/= | 114 (38.9) | 17 (35.4) |  |
|  | >500,000/= | 12 (4.1) | 6 (12.5) |  |
|  | <50,000/= | 127 (43.3) | 19 (39.6) |  |
| Dependent | No | 98 (33.4) | 14 (29.2) | 0.55 |
|  | Yes | 195 (66.6) | 34 (70.8) |  |
| Physical exercise | No | 32 (10.9) | 6 (12.5) | 0.70 |
|  | Yes | 261 (89.1) | 41 (85.5) |  |
| Smoking | No | 288 (98.3) | 47 (97.9) | 0.85 |
|  | Yes | 5 (1.7) | 1(2.1) |  |
| Alcohol consumption | No<br>Yes | 208 (71.0)<br>85 (29.0) | 37 (77.1)<br>11 (22.9) | 0.38 |
| Diet | Occasionally<br>Daily<br>Weekly<br>Monthly<br>Never | 5 (1.7)<br>245 (83.6)<br>37 (12.6)<br>5 (1.7)<br>1 (0.3) | 4 (8.4)<br>39 (81.3)<br>4 (8.3)<br>1 (2.1)<br>0 (0.0) | 0.102 |
| Diabetic history | No<br>Yes<br>I don't know | 188 (64.2)<br>96 (32.8)<br>9 (3.0) | 13 (27.1)<br>34 (70.9)<br>1 (2.0) | *<0.001 |
| Duration of ART | 1-10 years<br>11-20 years<br>21-37 years | 40 (13.7)<br>134 (45.7)<br>119 (40.6) | 3 (6.3)<br>13 (27.0)<br>32 (66.7) | *0.003 |

\*p value computed using Fisher exact test, ART: anti-retroviral therapy, TZs: Tanzania shillings, 1 USD = 2700 TZs

**Figure 1:**
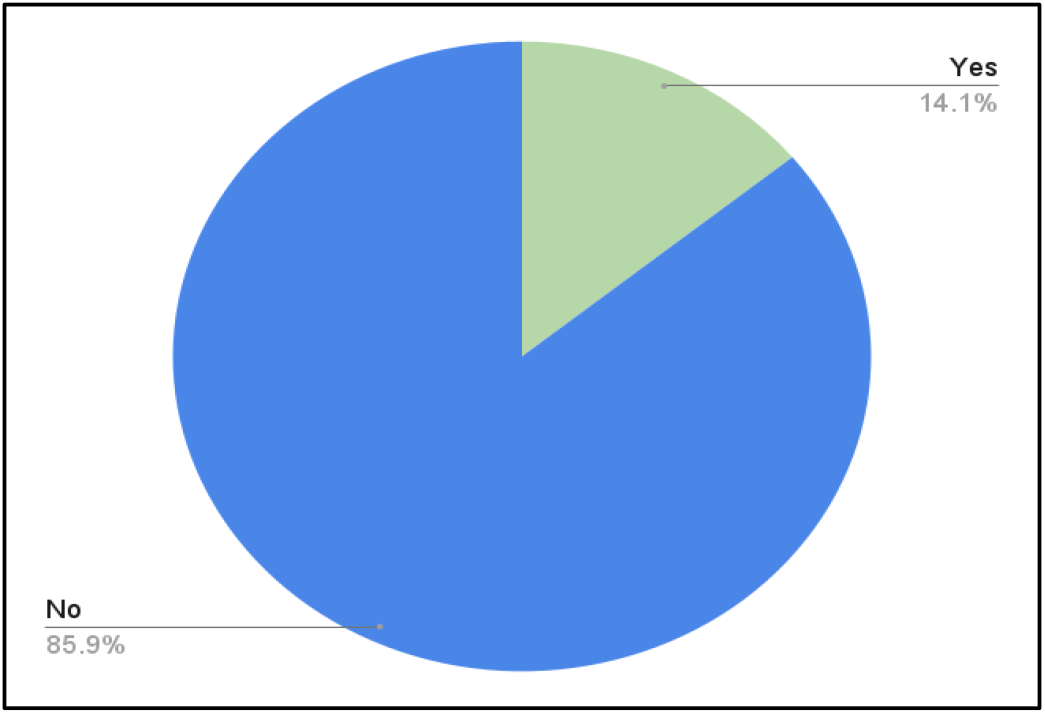
Prevalence of diabetes among HIV patients (n=341)

### Prevalence of high blood pressure among HIV patients

Out of 341 HIV patients, 23.5% (80) were hypertensive, Figure 2. The prevalence of hypertension was notably higher among individuals aged 36 to 62 years, with a rate of 71.3% (57), compared to other age groups p<0.001. Among hypertensive patients approximately 65% (52) were female. Most of the patients were dependants 81.3% (65).

**Figure 2:**
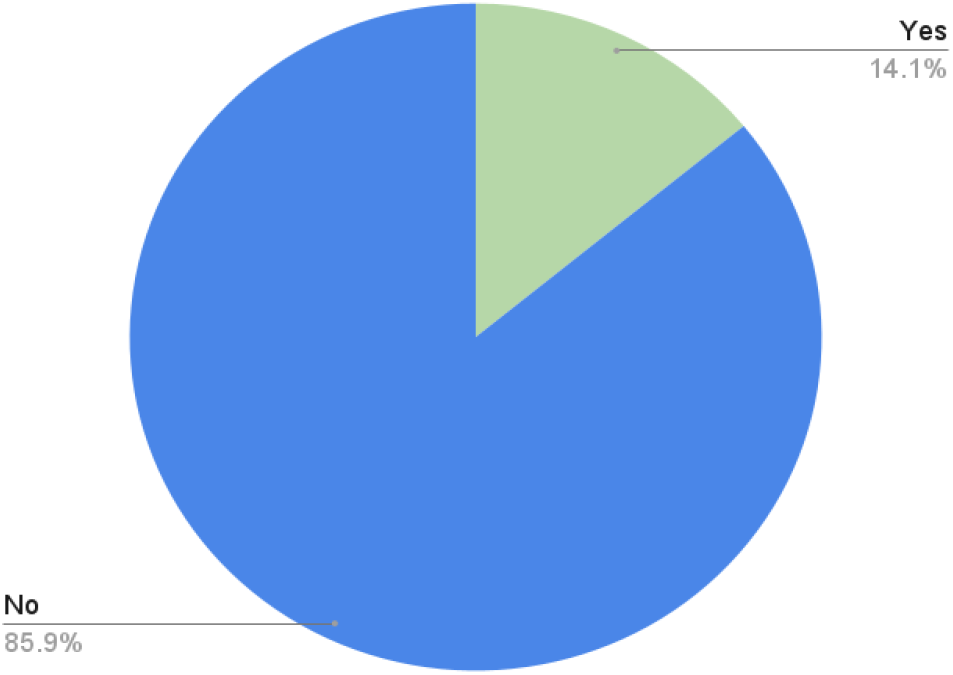
Prevalence of high blood pressure among HIV patients (n=341).

In terms of employment status, 63.8% (51) were self-employed, p=0.001, and 57.5% (46) had attained only a primary education level. A significant majority of the subjects had a family history of hypertension, accounting for 63.7% (51), p<0.001, Table 3.

**Table 3:** Association of Demographic characteristics with hypertension among HIV patients (n=341)

| variables |  | Blood Pressure status |  | P value |
| --- | --- | --- | --- | --- |
|  |  | Non-hypertensive n (%) | Hypertensive n (%) |  |
| Age | 8-35 | 93 (35.6) | 4 (5.0) | *<0.001 |
|  | 36-62 | 133 (51.0) | 57 (71.3) |  |
|  | >63 | 35 (13.4) | 19 (23.7) |  |
| Gender | Female | 175 (67.0) | 52 (65.0) | 0.73 |
|  | Male | 86 (33.0) | 28 (35.0) |  |
| Education level | No formal education | 6 (2.3) | 0 (0.0) | *0.008 |
|  | Primary education | 106 (40.6) | 46 (57.5) |  |
|  | Secondary education | 121 (46.4) | 22 (27.5) |  |
|  | Tertiary education | 28 (10.7) | 12 (15.0) |  |
| Employment status | Unemployed | 27 (10.3) | 4 (5.0) | *0.001 |
|  | Employed | 43 (16.5) | 15 (18.8) |  |
|  | Self employed | 127 (48.7) | 51 (63.8) |  |
|  | Student | 48 (18.4) | 1 (1.2) |  |
|  | Retired | 16 (6.1) | 9 (11.2) |  |
| Income in TZs | 200,000-500,000 | 33 (12.6) | 13 (16.3) | 0.03 |
|  | 50,000-200,000 | 97 (37.2) | 34 (42.5) |  |
|  | >500,000 | 10 (3.8) | 8 (10.0) |  |
|  | <50,000 | 121 (46.4) | 25 (31.2) |  |
| Dependent | No | 97 (37.2) | 15 (18.7) | 0.002 |
|  | Yes | 164 (62.8) | 65 (81.3) |  |
| Physical exercise | No | 30 (11.5) | 9 (11.2) | 0.73 |
|  | Yes | 231 (88.5) | 71 (88.8) |  |
| Smoking | No | 258 (98.9) | 77 (96.2) | 0.12 |
|  | Yes | 3 (1.1) | 3 (3.8) |  |
| Alcohol consumption | No | 194 (74.3) | 51 (63.7) | 0.06 |
|  | Yes | 67 (25.7) | 29 (36.3) |  |
| Diet | Occasionally | 5 (1.9) | 4 (5.0) | 0.28 |
|  | Daily | 220 (84.3) | 64 (80.0) |  |
|  | Weekly | 32 (12.3) | 9 (11.2) |  |
|  | Monthly | 3 (1.1) | 3 (3.3) |  |
| BP history | No | 167 (64.0) | 29 (36.3) | *<0.001 |
|  | Yes | 87 (33.3) | 51 (63.7) |  |
|  | I don't know | 7 (2.7) | 0 (0.0) |  |
| Duration of ART | 1-10 years | 37 (14.2) | 6 (7.5) | 0.28 |
|  | 11-20 years | 111 (42.5) | 36 (45.0) |  |
|  | 21-37 years | 113 (43.3) | 38 (47.5) |  |
| Diabetes | No | 233 (89.3) | 60 (75) | 0.001 |
|  | Yes | 28 (10.7) | 20 (25) |  |
| Salt intake | Daily | 215 (82.4) | 29 (36.2) | <0.001 |
|  | Weekly | 18 (6.9) | 7 (8.8) |  |
|  | Monthly | 28 (10.7) | 44 (55.0) |  |

\*p value computed using Fisher exact test, ART: anti-retroviral therapy, TZs: Tanzania shillings, 1 USD = Tanzania shillings

### Factors associated with diabetes among HIV patients

In bivariate logistic regression analysis, age, monthly income, diet and duration of HIV (set p-value <0.05) were included in multivariate logistic regression analysis. In the multivariate analysis, age, diet, income were significantly associated with diabetes mellitus. Those with age range 8-35 and 36-62 years were less likely to have diabetes as compared to those with greater than 63 years with AOR=0.017 (95% CI: 0.002-0.137), p<0.001, and (AOR=0.295 (95% CI: 0.137-0.636), p=0.002 respectively. Participants with income of TZs 500,000/= per month were more likely to develop diabetes as compared to other (AOR=4.112, 95% CI (1.199-14.108), p=0.025). Those who were taking fruit and vegetables were more likely to be diabetes as compare to other (AOR=0.15,95% CI (0.030-0.746), p= 0.03), Table 4.

**Table 4:** Bivariate and multivariate analyses of factors associated with diabetes in people living with HIV/AIDS receiving care at KCMC hospital.

| **Associated factors** | **N** | **N (%)**<br>**with**<br>**diabetes** | **COR (95%CI)** | **P**<br>**Value** | **AOR (95%CI)** | **P**<br>**Value** |
| --- | --- | --- | --- | --- | --- | --- |
| **Age(years)** |  |  |  |  |  |  |
| 8-35 | 97 | 1(2.1) | 0.02 (0.03-0.16) | <0.001 | 0.01 (0.002-0.13) | <0.001 |
| 36-62 | 190 | 29(60.4) | 0.36 (0.18-0.71) | 0.004 | 0.29 (0.13-0.63) | 0.002 |
| >63 | 54 | 18(37.5) | 1.0 |  | 1.0 |  |
| **Gender** |  |  |  |  |  |  |
| Female | 227 | 13(27.1) | 1.41 (0.71-2.79) | 0.31 |  |  |
| male | 114 | 35(72.9) | 1.0 |  |  |  |
| **Income in Tzs** |  |  |  |  |  |  |
| 200,000-500,000/= | 46 | 6(12.5) | 1.003 (0.38-2.68) | 0.99 | 4.11(1.19-14.10) | 0.02 |
| 50,000-200,000/= | 131 | 17(35.4) | 0.99 (0.49-2.01) | 0.99 |  |  |
| >500,000/= | 18 | 6(12.5) | 3.34 (1.12-9.96) | 0.03 |  |  |
| <50,000/= | 146 | 19(39.6) | 1.0 |  | 1.0 |  |
| **Physical Exercise** |  |  |  |  |  |  |
| No | 39 | 7(14.6) | 1.19 (0.47-3.03) | 0.71 |  |  |
| Yes | 302 | 41(85.4) | 1.0 |  |  |  |
| **Cigarette smoking** |  |  |  |  |  |  |
| No | 335 | 47(97.9) | 0.81 (0.09-7.14) | 0.85 |  |  |
| Yes | 6 | 1(2.1) | 1.0 |  |  |  |
| **Alcohol Consumption** |  |  |  |  |  |  |
| No | 96 | 37(77.1) | 1.37 (0.67-2.82) | 0.38 |  |  |
| Yes | 245 | 11(22.9) | 1.0 |  |  |  |
| **Diet** |  |  |  |  |  |  |
| Monthly | 7 | 1(2.1) | 0.21 (0.02-2.52) | 0.22 |  |  |
| Daily | 284 | 39(81.3) | 0.19 (0.05-0.77) | 0.02 | 0.15 (0.03-0.83) | 0.03 |
| Weekly | 41 | 4(8.3) | 0.13 (0.03-0.71) | 0.02 | 0.09 (0.01-0.62) | 0.01 |
| Occasionally | 9 | 4(8.3) | 1.0 |  | 1.0 |  |
| **Diabetic History** |  |  |  |  |  |  |
| No | 201 | 13(27.1) | 0.62 (0.07-5.29) | 0.66 |  |  |
| Yes | 130 | 34(70.8) | 3.18 (0.39-26.1) | 0.28 |  |  |
| I don't know | 10 | 1(2.1) | 1.0 |  |  |  |
| **Duration of HIV** |  |  |  |  |  |  |
| 1992-2002 | 54 | 18(37.5) | 4.59 (1.98-10.65) | <0.001 | 2.34 (0.88-6.19) | 0.08 |
| 2003-2013 | 115 | 19(39.6) | 1.11 (0.51-2.45) | 0.78 |  |  |
| 2014-2024 | 112 | 11(22.9) | 1.0 |  | 1.0 |  |

AOR = Adjusted odds ratio, COR = Crude odds ratio, TZs: Tanzania shillings, 1 USD = 2700 TZs

### Factors associated with hypertension among HIV patients

In bivariate logistic regression analysis, age, monthly income, dependable, hypertension history and salt intake (set p-value<0.05) were included in multivariate logistic regression analysis. In multivariate analysis age, income, hypertension history and salt intake were significantly associated with hypertension. Those with age range 8-35 years were less likely to have hypertension as compared to those with greater than 63 years with AOR=0.10 (95% CI: 0.03-0.41), p= 0.004, Participants with income of above TZs500,000/= per month were more likely to develop hypertension as compared to other (AOR= 3.50, 95% CI (0.91-13.49), p=0.03, Individuals who had not family history of hypertension were less likely to develop hypertension compared to those who had AOR=0.36 (95% CI: 0.19-0.68), p= 0.002 and those who were never taking salt were more likely to have hypertension compare to those took salt weekly with AOR= 5.73 (95% CI: 1.65-19.97), p= 0.006, Table 5.

**Table 5:** Bivariate and multivariate analyses of factors associated hypertension of people living with HIV/AIDS receiving care at KCMC hospital.

| Associated factors | N | N (%) with hypertension | COR (95%CI) | P Value | AOR (95%CI) | P value |
| --- | --- | --- | --- | --- | --- | --- |
| **Age(years)** |  |  |  |  |  |  |
| 8-35 | 97 | 4(5.0) | 0.07 (0.02-0.24) | <0.001 | 0.10 (0.03-0.41) | 0.004 |
| 36-62 | 190 | 57(71.25) | 0.07 (0.41-1.49) | 0.46 |  |  |
| >63 | 54 | 19(23.75) | 1.0 |  | 1.0 |  |
| **Gender** |  |  |  |  |  |  |
| Female | 227 | 52(65.0) | 0.91 (0.54-1.54) | 0.73 |  |  |
| Male | 114 | 28(35.0) | 1.0 |  |  |  |
| **Income in TZs** |  |  |  |  |  |  |
| 200,000-500,000/= | 46 | 13 (16.25) | 1.90 (0.88-4.13) | 0.10 |  |  |
| 50,000-200,000/= | 131 | 34 (42.5) | 1.69 (0.94-3.03) | 0.07 |  |  |
| >500,000/= | 18 | 8 (10.0) | 3.87 (1.39-10.78) | 0.01 | 3.50 (0.91-13.49) | 0.03 |
| <50,000/= | 146 | 25 (31.25) | 1.0 |  | 1.0 |  |
| **Physical Exercise** |  |  |  |  |  |  |
| No | 39 | 9 (11.25) | 0.89 (0.38-1.98) | 0.73 |  |  |
| Yes | 302 | 71 (88.75) | 1.0 |  |  |  |
| **Cigarette smoking** |  |  |  |  |  |  |
| No | 335 | 77 (96.25) | 0.29 (0.06-1.51) | 0.14 |  |  |
| Yes | 6 | 3 (3.75) | 1.0 |  |  |  |
| **Alcohol Consumption** |  |  |  |  |  |  |
| No | 96 | 51(63.75) | 0.61 (0.36-1.04) | 0.06 |  |  |
| Yes | 245 | 29(36.25) | 1.0 |  |  |  |
| **Diet** |  |  |  |  |  |  |
| Monthly | 7 | 3 (3.75) | 0.94 (0.13-6.90) | 0.95 |  |  |
| Daily | 284 | 64 (80) | 0.36 (0.09-1.39) | 0.14 |  |  |
| Weekly | 41 | 9 (11.25) | 0.35 (0.08-1.59) | 0.17 |  |  |
| Occasionally | 9 | 4 (5) | 1.0 |  |  |  |
| **Dependent** |  |  |  |  |  |  |
| No | 112 | 15 (18.75) | 0.39 (0.21-0.72) | 0.003 | 1.01 (0.45-2.22) | 0.97 |
| yes | 229 | 65 (81.3) | 1.0 |  | 1.0 |  |
| **Duration of ART** |  |  |  |  |  |  |
| 1-10 years | 43 | 6(7.5) | 0.48 (0.19-1.23) | 0.12 |  |  |
| 11-20 years | 147 | 36(45) | 0.96 (0.57-1.63) | 0.89 |  |  |
| 21-37 years | 151 | 38(47.5) | 1.0 |  |  |  |
| **Occupation** |  |  |  |  |  |  |
| Self employed | 178 | 51(63.8) | 2.71 (0.90-8.14) | 0.07 |  |  |
| Retired | 24 | 9(11.25) | 4.05 (1.06-15.4) | 0.04 | 2.97 (0.74-12.01) | 0.63 |
| Employed | 58 | 15(18.75) | 2.35 (0.71-7.44) | 0.16 |  |  |
| Student | 49 | 1(1.25) | 0.14 (0.02-1.32) | 0.08 |  |  |
| Unemployed | 31 | 4(5) | 1.0 |  | 1.0 |  |
| **Hypertension history** |  |  |  |  |  |  |
| No | 203 | 29(36.25) | 0.28 (0.16-0.48) | <0.001 | 0.36 (0.19-0.68) | 0.002 |
| Yes | 138 | 51(63.75) | 1.0 |  | 1.0 |  |
| **Salt intake** |  |  |  |  |  |  |
| Never | 38 | 26(32.5) | 5.57 (1.84-16.9) | 0.002 | 5.73 (1.65-19.97) | 0.006 |
| Monthly | 34 | 18(22.5) | 2.89 (0.96-8.71) | 0.05 | 0.00 (0-0.001) | 1.0 |
| Daily | 244 | 29(36.3) | 0.35 (0.13-0.90) | 0.03 | 0.49 (0.17-1.42) | 0.19 |
| Weekly | 25 | 7(8.7) | 1.0 |  | 1.0 |  |

AOR = Adjusted odds ratio, COR = Crude odds ratio, ART: anti-retroviral therapy
1 USD = 2700 TZs, TZs: Tanzania shillings

## DISCUSSION

Diabetes and hypertension among HIV patients and their associated factors are the global health challenges. This study aimed to determine prevalence and associated factors for diabetes and high blood pressure among HIV patients at KCMC hospital in Moshi district. The overall prevalence of diabetes among HIV patients was 14.1% which is higher than previously reported in Tanzania which was 13% in in Mwanza (Jeremiah *et al*., 2020b) and 8.8% in Ethiopia (Manavalan et al., 2022). In other hand our findings is lower than the study conducted in Uganda which was 4.7% (Manavalan *et al*., 2022). The observed differences in the prevalence between our findings and previous studies could be because of low sample size. High prevalence of diabetes among HIV patients implies that development of non-communicable diseases still to be a health challenge, so clinician should implement diabetes screening and treatments program among HIV patients.

Additionally, our study reports high prevalence of diabetes among older age group above or equal to 63 years. This implies that individuals with older age have predisposing factors that could explain the higher prevalence including decreasing of physical activity level and duration of ART (Nagai *et al*., 2023). Also our study report high prevalence of diabetes among self-employed may be they have higher income which could have lead them to have sedentary life style (Duguma *et al*., 2020). Study report high prevalence of diabetes among HIV patients who had diabetic family history because diabetes can be inherited from one generation to another (Hertz *et al*., 2022). The prevalence of diabetes in our study reported to be higher among patients who were in long duration of ART for 21 to 37 years which could lead to interference of the body glucose metabolism as highlighted in a study conducted in south Africa (Manne-Goehler *et al*., 2019).

The predetermined risk factors for diabetes among PLWHIV in this study were older age, High income, and Daily intake of diet consists fruits and vegetables while it was different in the study conducted in Ethiopia which showed long duration of ART, alcohol consumption and physical exercise to be an associated factors (Manavalan *et al*., 2020).

The overall prevalence of hypertension among HIV patients was 23.5% which was higher than previously reported in Tanzania, one from northern Tanzania which was 5.5% (Manavalan *et al*., 2020). Difference in prevalence could be due to variation of sample size and geographical locations, and other conducted in Dar es salaam 12.5% (Marina et al., 2016).

In other hand our findings is lower than the study conducted which was 28.5% in Kenya (wambula et al., 2021). Additionally, our study reports high prevalence of hypertension among older age group between 36-62 years. This implies that individuals with older age have predisposing factors that could explain the higher prevalence including decreasing of physical activity and dietary patterns (Moyo-Chilufya *et al*., 2023). Also our study report high prevalence of hypertension among primary education individuals due to the low awareness of hypertension, a low hypertension awareness among PLHIV indicates a missed opportunity to screen and diagnose hypertension (Isaac Derick and Khan, no date).The prevalence of hypertension in our study reported to be higher among patients who were self-employed due to stressful which can exacerbate health issue (Mwakyandile et al., 2024).

The prevalence of hypertension in our study reported to be higher among patients who had income between 50,000-200000 TZs because of their income they might be aware of their health condition due to regular check-ups, leading to higher reported prevalence of hypertension (Davis *et al*., 2021). Also prevalence of hypertension in our study reported to be higher among patients who were dependable due to high workload because reliable individuals may have demanding job or role that can be stressful and contribute to hypertension, the pressure of meeting high expectation or managing multiple responsibility can affect blood pressure (Tegegne et al., 2023). Study report high prevalence of hypertension among HIV patients who had hypertension family history because hypertension can be inherited from one generation to another, these genetic factors are independent of HIV but can interact with the virus and its treatment (Harimenshi et al., 2022).

Also, the prevalence of hypertension is higher in people with diabetes, because diabetes might compromise kidney function leading to diabetic nephropathy, which affect the kidney’s ability to regulate blood pressure. Combine with HIV-related kidney complications, this can further increase hypertension risk (Hertz *et al*., 2022). The people who were monthly taking salts food were found to have higher prevalence of hypertension due to their hypertensive condition, because excessive salt intake could lead to high blood pressure. The predetermined risk factors for hypertension among PLWH in this study were age, income, and hypertension family history while the associated factor in the studies conducted in Kenya were sex, BMI and history of alcohol consumption (Mbuthia, Magutah and McGarvey, 2021).

## Strengths and Limitations of the study

The study highlights a significant prevalence of diabetes among individual living with HIV, which is critical for understand the burden of comorbidities in Tanzania population. These finding suggest socioeconomic factors play a significant role in the prevalence of diabetes among PLWH, offering insights into potential areas for targeted intervention. Utilizing multivariable regression analyses allows for an insightful understanding of the association between HIV and these comorbidities. This methodological rigor enhances the credibility of the findings.

The cross-sectional nature of the studies precludes establishing temporality and causality between HIV status and diabetes/hypertension prevalence. Longitudinal studies are needed to better understand the incidence and progression of these condition in relation to HIV infection over time.

## Conclusion

The result suggests that social determinant, rather than HIV status alone, may significantly influence the prevalence of diabetes and hypertension. This insight can inform public health strategies and resource allocation, emphasizing the need to address broader socio-economic factors in managing health outcomes for people living with HIV. More research should be done with cohort nature in order to establish the association of diabetes and hypertension in PLWH. Healthcare givers should be made aware of the problem so as to be able to identify it early investigate and give a promptly management. Implement regular screening of diabetes and hypertension among HIV patients. Educate patients about the risks of diabetes and hypertension in the context of HIV based on the life style which can help in management and prevention.

## Data Availability

All relevant data are within the manuscript and its Supporting Information files.

## Acknowledgements

The authors would like to thank the management of KCMC Hospital permission to conduct the study.

## Authors’ contributions

DCK conceived the study, analyzed the data, and drafted the manuscript, ALH, GF, JB and EM performed all the measurement and revised the manuscript, DM,BMN, JJS, FM, IL, contributed to interpretation of the data and critical review of the manuscript. All authors read and approved the final manuscript.

## Data availability

The datasets used and/or analyzed during the current study are available from the corresponding author on reasonable request.

## Consent for publication

NA

